# Macular Structure Characteristics in Unilateral Idiopathic Full-Thickness Macular Hole and the Healthy Fellow Eyes

**DOI:** 10.1101/2024.03.16.24304402

**Authors:** Yi-Ting Hou, Chung-May Yang, Yi-Ting Hsieh

## Abstract

**Aims:** To investigate the characteristics of the macular structure and foveal pit in eyes with lesions and healthy fellow eyes of patients with unilateral full-thickness macular holes (FTMH).

**Methods:** Patients with unilateral FTMH were retrospectively enrolled as the study group, and their age- and sex-matched individuals with no vitreomacular diseases as the control group in a medical center. FTMHs were classified as FTMH with lamellar hole-associated epiretinal proliferation (LHEP), FTMH without LHEP or FTMH without vitreomacular separation. Macular structure parameters, including foveal base width (FBW), central foveolar thickness (CFT), central subfield thickness (CST), central subfield volume (CSV), and retinal artery trajectory (RAT), were measured using optical coherence tomography and fundus photography. These parameters were compared among different FTMH groups.

**Results:** A total of 68 patients (39 women) with unilateral FTMH and 68 healthy controls were enrolled. The RAT of the lesioned eyes (0.19±0.06) and the healthy fellow eyes (0.14±0.04) were both smaller than those of the normal controls (0.37±0.14) (*P*<0.001 for both). The FBW of the healthy fellow eyes (446.8±98.2 µm) were significantly larger than those in the control group (338.4±80.6 µm, *P*<0.001). No significant differences in any macular parameters were noted among the three different types of FTMH.

**Conclusions:** Patients with unilateral FTMH had a wider RAT in both the lesioned and healthy eyes, and a wider foveal base in their healthy fellow eyes than in normal controls. Such macular structure characteristics may be prone to macular hole formation.

## INTRODUCTION

Full-thickness macular hole (FTMH) predominantly affects the aging population without significant underlying disease or etiologies.^1,2^ The prevalence is higher in females than males.^3^ Patients with a full-thickness macular hole would suffer from visual acuity deterioration if remained untreated.^4–6^ Although the exact pathogenesis of FTMH remains unclear, the most commonly accepted mechanism is the traction force at the vitreoretinal interface, which would induce foveal deformity and tissue disruption.^2^ Traditionally, FTMH was thought to be caused mainly by the anteroposterior vitreomacular traction from the partially detached posterior hyaloid, and Gass proposed the classic 4-stage concept of FTMH formation.^7,8^ With the recent advances in optical coherence tomography (OCT), the macular structure and its relationship with posterior hyaloid in FTMH can be identified and quantified more precisely.^9,10^ Recent studies have found that traction from epiretinal membrane (ERM) may also give rise to the development of FTMH in eyes with posterior vitreous detachment (PVD).^6,11,12^ Furthermore, our study team also found that FTMH could develop in eyes without vitreomacular separation (VMS); some of them had ERM, while some did not.^13^ This suggests that the force underlying FTMH may also come from the centrifugal tangential traction of the posterior hyaloid or ERM.

In our previous study, we identified a wide-based foveal pit as a specific foveal pattern that was associated with higher incidences of vitreomacular abnormalities, including ERM and FTMH.^14^ We proposed that the widening of the foveal pit may be related to tangential centrifugal traction at the macula, which may keep dragging the macula centrifugally, leading to some vitreomacular interface change including internal limiting membrane (ILM) cleft formation, glial cell proliferation, and eventually ERM formation; on the other hand, such traction force may lead to foveal thinning, lamellar macular hole formation, and FMTH formation in the long term. Sometimes ERM and MH may exist simultaneously. Another study from our team has demonstrated that fellow eyes with unilateral ERM had a larger foveal base width (FBW) and a wider retinal artery trajectory (RAT) than the normal population;^14,15^ These findings support our hypothesis that some tangential centrifugal traction may cause the development of ERM due to glial cell proliferation after the formation of the ILM cleft. We aimed to elucidate whether tangential centrifugal traction is also a possible cause for the development of FTMH. It is well known that patients with unilateral FTMH also have an increased risk of developing FTMH in their fellow eyes.^16–18^ Previous studies also demonstrated that symmetric foveal configuration exists in both eyes.^19,20^ Since the macular structure changes after the formation of FTMH, the macular structure of the fellow eyes of patients with unilateral FTMH should also provide some information regarding the vitreoretinal interface and macular structure, that may result in the formation of FTMH. In this study, we investigated the macular structure parameters in the lesioned eyes as well as the asymptomatic fellow eyes of patients with unilateral FTMH and compared them with normal controls as well as among different types of FTMH.

## METHODS

### Study subjects

This retrospective observational study adhered to the tenets of the Declaration of Helsinki and was approved by the Research Ethics Committee of National Taiwan University Hospital (No.202111031RINA). The requirement for informed consent was waived owing to the retrospective nature of the study. Patients who presented with unilateral idiopathic FTMH and had undergone macular OCT examinations under Dr. YT Hsieh or Dr. CM Yang at the National Taiwan University Hospital from January 2018 to December 2021 were retrospectively enrolled as the study group. Exclusion criteria included the presence of high myopia (spherical equivalent of -6.0 diopters or less or an axial length greater than 26 mm), posterior staphyloma, history of retinal detachment, diabetic retinopathy, retinopathy of prematurity, retinal vascular diseases, retinal dystrophy, choroidal neovascularization, posterior uveitis, previous intraocular surgeries except for cataract surgery, and any condition that may cause secondary FTMH formation. Because one specific type of FTMH, macular hole without VMS, was less prevalent, we also recruited cases of this type that had been enrolled in our previous study^13^ for analysis in the present study. During the same period, healthy normal controls were recruited from patients who visited our hospital for routine examinations, including macular OCT for cataract surgery or refractive surgery, and had no apparent vitreoretinal diseases in either eye. Those who were sex- and age-matched (same sex, age difference ≤ 5 years) to the cases in the study group were chosen as the control group. Only one eye was randomly selected from each participant in the control group for analysis.

For each patient, comprehensive history taking (including age, sex, and past systemic and ocular history) and ocular examination (including best-corrected visual acuity, slit-lamp evaluation, and fundoscopy) were performed. The axial length was measured using laser interferometric biometry (Lenstar LS900; HAAG-STREIT AG, Bern, Switzerland). All patients underwent macular B-scan OCT and en face OCT angiography using the Optovue Avanti RTVue XR OCT (Optovue, Inc., Fremont, CA, USA). Fundus images were obtained using color fundus photography, fundus autofluorescence, or infrared imaging. Patients with incomplete data were excluded from this study.

### Macular structure parameters

The macular structural parameters measured in this study included FBW, central foveolar thickness (CFT), central subfield thickness (CST), central subfield volume (CSV), and RAT. The measurement of these parameters has been described previously,^15^ and will be mentioned briefly here. FBW was defined as the distance between two intersections of the ILM and a line parallel to the underlying retinal pigment epithelium layer 10 µm above the lowest point of the foveal pit in the B-scan OCT image. We calculated the vertical and horizontal sections of the B-scan OCT image and recorded the mean value as the FBW. In the study group, only fellow eyes with normal macular structures and foveal contours were evaluated for FBW. CFT was measured as the distance between the lowest point of the pit and the base of the retinal pigment epithelium layer. CST and CSV were measured at the central 1 mm-diameter area around the foveal pit, and both data were retrieved automatically using the software Optovue Avanti RTVue after the centration was checked manually for precision. CST is the average retinal thickness over this area, and CSV is the total volume of the retina in this area. The technique for calculating RAT was modified according to the method proposed by Yoshihara et al,^21^ and has been described in our previous study. We first rotated the fundus images either clockwise by 90 degrees in the right eye or counterclockwise by 90 degrees in the left eye using ImageJ (ImageJ version 1.47; National Institutes of Health, Bethesda, MD). Second, we manually dotted the arcade arteries originating from the optic disc, and 24 dots were labeled in each fundus photograph. The X-Y coordination was calculated using ImageJ software, and the coordinated data were fitted to the best curve with a second-degree polynomial equation ax2/100 + bx + c. Constant “a” represents the slope of the main arcade morphology, where the smaller “a” makes the artery curve far from the macula, and the larger “a” means the curve is closer to the macula.

### Statistical analysis

The macular structure parameters were compared between eyes with FTMH lesions and normal controls, as well as between fellow eyes with FTMH and normal controls. Continuous variables were analyzed using Student’s *t*-tests, and categorical variables were examined using the chi-square test or Fisher’s exact test. We also further divided FTMH into three different types: (1) Gass’ stage 2 to 4 FTMH without lamellar hole-associated epiretinal proliferation (LHEP), (2) Gass’ stage 2 to 4 FTMH with LHEP, and (3) FTMH without VMS, and compared the macular structure parameters among them. Univariate and multivariate logistic regression analyses were performed to investigate predisposing factors for FTMH development. Receiver operating characteristic (ROC) curves were plotted using FBW and RAT to predict whether the fellow eye had FTMH, and the areas under the curve (AUCs) were calculated. Statistical significance was set at *P* < 0.05. Statistical analyses were performed using IBM SPSS Statistics for Windows version 25 (IBM Corp., Armonk, NY, USA).

## RESULTS

### Demographic data

This study recruited 68 patients in each of the unilateral FTMH study groups and the age- and sex-matched control group. Both groups comprised 29 males and 39 females. The average age was 60.7 ± 8.7 years in the study group and 62.0 ± 13.5 years in the control group (*P* = 0.83). The average axial length was 23.75 ± 1.25 mm in the unilateral FTMH group and 23.73±0.94 mm in the age and sex-matched control group (*P* = 0.82).

### Macular structure parameters in unilateral FTMH and normal controls

Table 1 compares macular structure parameters among the lesioned eyes of patients with unilateral FTMH, fellow eyes of patients with unilateral FTMH, and normal control eyes. The mean FBW was 446.8 ± 98.2 µm in the fellow eye of patients with unilateral FTMH, which was much higher than that in the control group (338.4 ± 80.6 µm, *P* < 0.001). The mean RAT was 0.19 ± 0.06 in the lesioned eye and 0.16 ± 0.04 in the fellow eye of the patients with unilateral FTMH, both smaller than the mean RAT in the control group (0.37 ± 0.14, *P* < 0.001 for both). This suggests that the RAT was wider in the lesioned eyes as well as in the fellow eyes of patients with unilateral FTMH than in normal controls. No significant differences in RAT were present between the lesioned eyes and fellow eyes of patients with unilateral FTMH (P = 0.248). As for CFT, CST, and CSV, no significant differences existed between the fellow eyes with unilateral FTMH and the normal control eyes (*P* > 0.05).

**Table 1.** Macular structure parameters in the lesioned eyes of unilateral full-thickness macular hole, contralateral unaffected eyes, and matched disease-free control eyes.

|  | FTMH |  | Fellow |  | Control |  | *P* value |  |  |
| --- | --- | --- | --- | --- | --- | --- | --- | --- | --- |
|  | N=68 |  | N=68 |  | N=68 |  | FTMH vs. Fellow | FTMH vs. Control | Fellow vs. Control |
|  | Mean | SD | Mean | SD | Mean | SD |  |  |  |
| AXL (mm) | 23.75 | 1.25 | 23.71 | 1.02 | 23.73 | 0.94 | 0.82 | 0.38 | 0.24 |
| FBW (μm) | - | - | 446.8 | 98.2 | 338.4 | 80.6 | - | - | **<0.001** |
| CFT (μm) | - | - | 201.7 | 32.7 | 203.2 | 25.0 | - | - | 0.16 |
| CST (μm) | - | - | 245.5 | 35.3 | 248.2 | 17.2 | - | - | 0.61 |
| CSV (μm<sup>3</sup>) | - | - | 0.19 | 0.03 | 0.19 | 0.02 | - | - | 0.43 |
| RAT | 0.19 | 0.06 | 0.16 | 0.04 | 0.37 | 0.14 | 0.25 | **<0.001** | **<0.001** |

A value of $P < 0.05$ was taken as statistically significant.
FTMH= Full-thickness macular hole, AXL=Axial length, FBW=Foveal base width, CFT=Central foveal thickness, CST=Central subfield thickness,
CSV=Central subfield volume, RAT=Retinal artery trajectory, SD=Standard deviation

#### Comparison of Macular anatomical structures among different types of FTMH

We divided FTMH into three groups based on different characteristics and vitreomacular association: FTMH without LHEP. FTMH with LHEP and FTMH without VMS. The FTMH size of the affected eye and the macular structure characteristics, including FBW, CFT, CST, CSV, and RAT of the fellow eye, were compared among the three groups. As shown in Table 2, there were no significant differences among the three groups in any macular parameter.

**Table 2.** Macular structure parameters in fellow eyes of the unilateral full-thickness macular hole with different patterns.

|  | MH without LHEP |  | FTMH with LHEP |  | FTMH without VMS |  | *P* value |
| --- | --- | --- | --- | --- | --- | --- | --- |
|  | N=37 |  | N=11 |  | N=20 |  |  |
|  | M | SD | M | SD | M | SD |  |
| FTMH size (μm) | 439.7 | 233.8 | 444.4 | 195.0 | 392.6 | 324.8 | 0.80 |
| FBW (μm) | 450.3 | 105.6 | 446.5 | 114.6 | 432.9 | 65.0 | 0.85 |
| CFT (μm) | 203.4 | 29.1 | 219.7 | 31.7 | 221.5 | 39.2 | 0.15 |
| CST (μm) | 238.9 | 37.7 | 260.7 | 24.5 | 248.9 | 35.3 | 0.18 |
| CSV (μm<sup>3</sup>) | 0.19 | 0.03 | 0.20 | 0.17 | 0.19 | 0.03 | 0.40 |
| RAT | 0.16 | 0.04 | 0.16 | 0.28 | 0.17 | 0.07 | 0.82 |

A value of $P < 0.05$ was taken as statistically significant.
FTMH=Full-thickness macular Hole, LHEP=Lamellar hole-associated epiretinal proliferation, VMS=Vitreomacular separation, AXL=Axial length, FBW=Foveal base width, CFT=Central foveal thickness, CST=Central subfield thickness, CSV=Central subfield volume, RAT=Retinal artery trajectory, M=Mean, SD=Standard deviation

### Sexual differences in Macular structure characteristics

Table 3 shows the differences in macular structure parameters between males and females in the age-matched control and study groups. In the age-matched control group, the female had larger FBW (370.1 ± 88.5 vs. 294.1 ± 37.1 µm, *P* < 0.001), thinner CFT (198.3 ± 18.7 vs. 213.4 ± 34.3 µm, *P* = 0.023), thinner CST (244.6 ± 17.8 vs. 253.5 ± 14.1 µm, P=0.016), smaller CSV (0.19 ± 0.01 vs. 0.21 ± 0.02 µm, *P* = 0.004) and wider RAT (0.35 ± 0.14 vs. 0.41 ± 0.13 µm, *P* = 0.030) than males. In the study group, females also had larger FBW and thinner CFT than males. There were no statistically significant differences in CST, CSV, or RAT scores between males and females in the study group. Interestingly, we found that RAT was larger in females than males in the normal population, but RAT in males in the study group was wider than in males in the normal group and females in the study group; however, the difference was not statistically significant (*P* = 0.275).

**Table 3.** Comparison of macular structure parameters between males and females in normal subjects and in the fellow eyes of unilateral full-thickness macular hole.

|  | Normal controls |  |  | Fellow eyes of unilateral FTMH |  |  |
| --- | --- | --- | --- | --- | --- | --- |
|  | Male<br>(n=29) | Female<br>(n=39) | P value | Male (n=29) | Female<br>(n=39) | *P* value |
| Age (years) | 60.4±16.3 | 62.1±9.56 | 0.15 | 61.69±9.18 | 60.00±8.22 | 0.428 |
| FBW (μm) | 294.1±37.1 | 370.1±88.5 | **<0.001** | 417.1±87.5 | 465±90.3 | **0.032** |
| CFT (μm) | 213.4±34.3 | 198.3±18.7 | **0.023** | 219.60±34.5 | 204.63±30.5 | **0.045** |
| CST (μm) | 253.5±14.1 | 244.6±17.8 | **0.016** | 250.51±34.4 | 241.5±35.9 | 0.325 |
| CSV (μm<sup>3</sup>) | 0.21±0.02 | 0.19±0.01 | **0.004** | 0.2±0.03 | 0.19±0.03 | 0.191 |
| RAT | 0.41±0.13 | 0.35±0.14 | **0.030** | 0.15±0.04 | 0.17±0.06 | 0.275 |

A value of $P < 0.05$ was taken as statistically significant.
FTMH=Full-thickness macular Hole, AXL=Axial length, FBW=Foveal base width, CFT=Central foveal thickness, CST=Central subfield thickness, CSV=Central subfield volume, RAT=Retinal artery trajectory

### Predicting FTMH with OCT parameters of the Fellow Eyes

We used a logistic regression model to investigate predisposing factors for FTMH. In the univariate analysis, a wider FBW (odds ratio [OR] = 1.014, 95% confidence interval [CI] = 1.008–1.020, *P* < 0.001) and wider RAT (OR = 0.793, per 0.01, 95% CI = 0.735–0.862, *P* < 0.001) were associated with FTMH in fellow eyes. After adjustment for age, sex, and axial length in multivariate analysis, FBW (OR = 1.021, 95% CI = 1.010–1.033, *P* < 0.001) and RAT (OR = 0.749, per 0.01, 95% CI = 0.655–0.853, *P* < 0.001) remained significantly associated with the diagnosis of FTMH (Table 4).

**Table 4.** Logistic regression analysis for the predisposing factors associated with full-thickness macular hole.

|  | Simple regression |  |  | Multiple regression |  |  |
| --- | --- | --- | --- | --- | --- | --- |
|  | Odds ratio | 95% CI | *P* value | Odds ratio | 95% CI | *P* value |
| Age (year) | 0.997 | 0.964–1.031 | 0.86 | 0.971 | 0.888–1.062 | 0.53 |
| Sex |  |  |  |  |  |  |
| Female | 1 (Reference) |  |  | 1 (Reference) |  |  |
| Male | 0.856 | 0.438–1.673 | 0.65 | 0.275 | 0.049–1.528 | 0.14 |
| AxL (mm) | 1.237 | 0.832–1.839 | 0.24 | 1.073 | 0.539–2.138 | 0.84 |
| FBW (μm) | 1.014 | 1.008–1.020 | **<0.001** | 1.021 | 1.010–1.033 | **<0.001** |
| CFT (μm) | 1.009 | 0.996–1.022 | 0.16 |  |  |  |
| CST (μm) | 0.997 | 0.984–1.010 | 0.62 |  |  |  |
| CSV (μm<sup>3</sup>) | 0.002 | 0.001–6554.66 | 0.42 |  |  |  |
| RAT (per 0.01) | 0.793 | 0.735–0.862 | **<0.001** | 0.749 | 0.655–0.853 | **<0.001** |

Data presented as mean ± standard deviation. A value of $P < 0.05$ was taken as statistically significant.
AxL=Axial length, FBW=Foveal base width, CFT=Central foveal thickness, CST=Central subfield thickness, CSV=Central subfield volume, RAT=Retinal artery trajectory

We subsequently plotted ROC curves using FBW and RAT to predict whether the fellow eye had FTMH, and the AUCs were 0.874 and 0.903 for FBW and RAT, respectively. The best threshold for FBW was 381 µm, with a sensitivity of 82.1% and specificity of 89.8%. The best threshold for RAT was 0.215, with a sensitivity of 92.1% and a specificity of 79.6%. (Figure 1)

**Figure 1:**
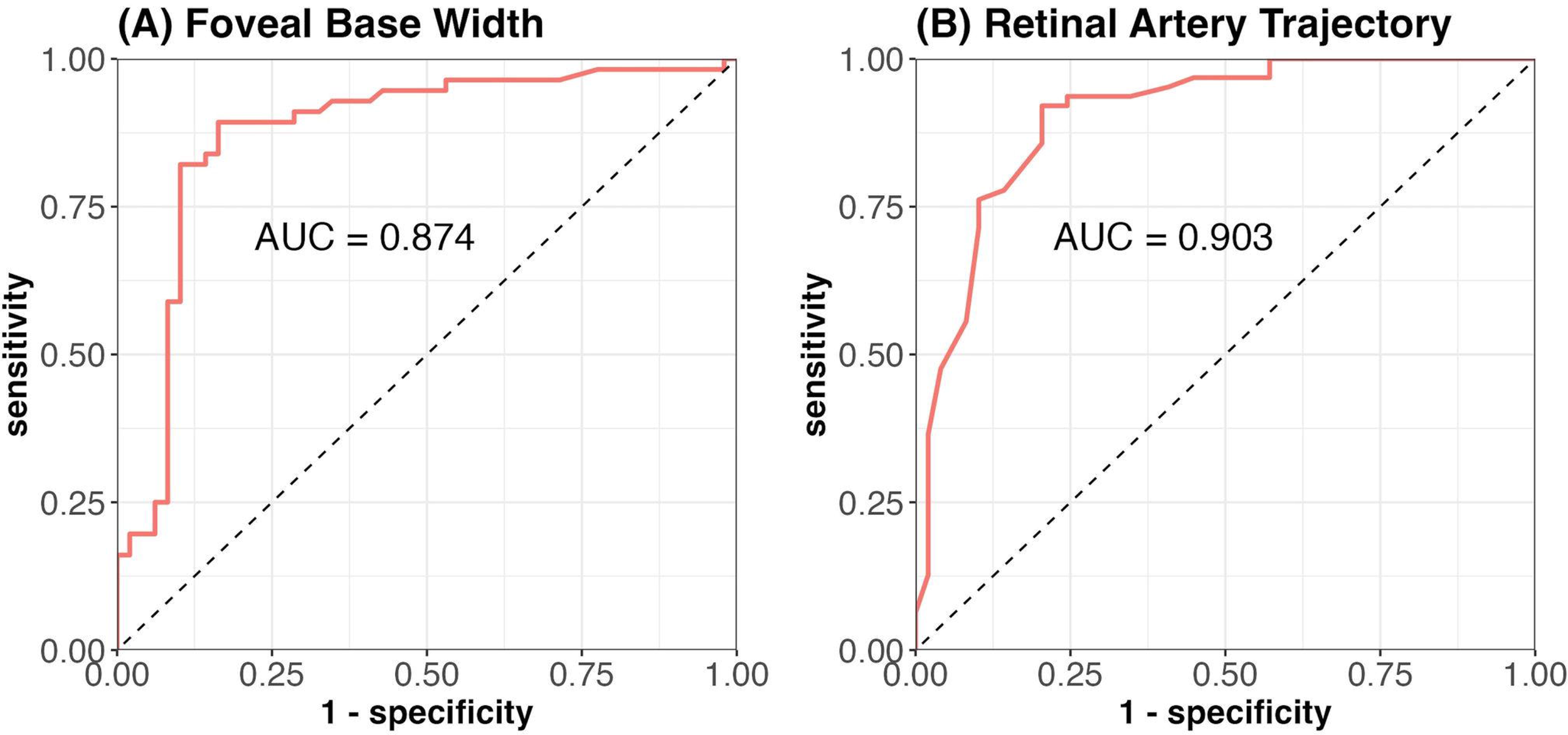
Receiver operation characteristic curves for predicting the presence of an idiopathic macular hole using the foveal base width (FBW) or retinal artery trajectory (RAT) of the fellow eye. (A) For FBW, the area under the curve was 0.874, and the best threshold of FBW was 381.0 μm. (B) For RAT, the area under the curve was 0.903, and the best threshold of RAT was 0.215.

## DISCUSSION

It is well known that patients with unilateral MH would have a higher risk of developing FTMH in their fellow eye.^4,16,18^ This raised our interest in studying whether predisposing factors for FTMH are also present in the fellow eyes of patients with unilateral FTMH. Through OCT, early changes in foveal configuration and visualization of the vitreomacular relationship could provide more information about the pathogenesis of FTMH formation. Two major mechanisms, anteroposterior vitreous traction and tangential vitreous traction (tangential traction), over the foveal area, are generally accepted as the main contributors to FTMH formation.^6,8^ However, the exact pathogenesis remains unclear. In the current study, we found that the lesioned eyes and fellow eyes of patients with unilateral MH had a wider foveal base and a wider RAT than age-matched healthy controls. This implies that increasing centrifugal traction parallel to the retina may exist in these patients, which results in the widening of the RAT and foveal base and the subsequent formation of MH in one eye. Yoshihara et al.^21^ found that in patients with unilateral FTMH, the RAT was wider in the FTMH eyes than in the healthy fellow eyes, and they also proposed that tangential traction may first widen the main arcades of retinal vessels and then induce larger traction over the central foveal area to induce the formation of FTMH. Although this result was different from what we found in the present study, they also found that the RAT of FTMH eyes was highly correlated with that of their healthy fellow eyes. Both results indicated that RAT was significantly associated with the formation of FTMH. Because the vitreous is usually attached tightly to retinal vessels, the larger distance between vessels may exacerbate the tangential force on the vitreomacular interface and the central macula, which also widens the foveal base.^14,15,21,22^ Additionally, we also found that the CST and CFT were thinner in the fellow eye of patients with unilateral FTMH than in healthy controls, although no significant statistical differences were noted. These results were similar to those of Kumagai et al.^23^ This is compatible with our hypothesis that tangential traction plays a vital role in FTMH formation with stretching of the central retina. Furthermore, no significant FBW and RAT differences were found in fellow eyes of unilateral FTMH eyes among all three groups, including the FTMH without LHEP, FTMH with LHEP, and FTMH without VMS groups. All three groups had wider RAT and FBW. Previously, our study group reported that the strong tangential traction thickened posterior hyaloid without VMS has a different pathomechanism of FTMH formation, which had no anteroposterior traction from conventional VMT.^13^ Our results from this study support that eyes with typical idiopathic FTMH may share similar macular structure characteristics with eyes with FTMH without VMS. We propose that continuous tangential traction from the posterior hyaloid or ILM may disrupt the ILM, make the retina at the central fovea more fragile, develop cystic changes in the foveal wall, induce lamellar macular hole and LHEP formation, and finally cause _FTMH._6,13,15

Many studies have reported that macular parameters are influenced by multiple variables,^24^ such as age,^25^ axial length,^26^ and sex.^27^ Therefore, we included these factors in the univariate and multiple regression models. Only FBW and RAT were found to be associated with the development of FTMH. Additionally, no significant differences existed in the RAT between the affected and fellow eyes in patients with unilateral MH. Both ROC curves using the FBW and RAT of the fellow eyes showed good predictions in the presence of idiopathic FTMH. Therefore, we believe that both a wider FBW and RAT are anatomic predisposing factors for FTMH rather than a post-MH change.

To better understand the macular structural differences between males and females, we compared macular parameters in terms of sexual differences. Our previous studies have reported that females have a wider FBW and a wider RAT than males in healthy eyes.^15^ In the current study, we found that females had wider FBW, thinner CFT, CST, CSV, and wider RAT compared to the male in normal controls. These results are consistent with those of several previous studies,^14,15,21^ which indicated that females might have a firmer centrifugal tangential force over the foveola area, making the central retina more fragile and thinner.^15^ CFT is the thickness at the center of the foveola; widening the foveal area would contribute to a thinner umbo of foveola and the tendency for MH formation. This phenomenon can also explain the higher incidence of MH and ERM in females than in men. In some population-based studies, females were 2.2–3.3 times more likely to be affected than males by the development of FTMH.^3,28^ In the unilateral FTMH study group, however, we found that the RAT showed not only no statistical differences between male and female, but also that the males had even wider RAT than female population. The results indicated that stronger tangential traction might be needed for males for the development of FTMH.

Our study had several limitations. First, this study was designed as a cross-sectional and retrospective data collection study. Second, it is not certain whether FTMH will develop in the fellow eyes that we studied. However, we sought to reduce the bias and statistical errors by matching age and sex to those in the study group. Thus, our findings are accurate, valuable, and clinically applicable.

In conclusion, we found that eyes with FTMH had a wider RAT than age- and sex-matched normal controls. As for the healthy fellow eyes with unilateral FTMH, the FBW was wider, and the RAT was wider than that of normal controls. A wider FBW and a wider RAT in healthy eyes were predictive of the presence of FTMH in the contralateral eyes. This suggests that strong tangential traction may contribute to FTMH development by dragging the foveal pit and thinning the central foveola.

Females had wider FBW, wider RAT, thinner CST, CFT, and CSV than males, which can explain the higher prevalence of MH in females.

## Data Availability

All data produced in the present study are available upon reasonable request to the authors.

## References

1. Yeh P-T, Chen T-C, Yang C-H, Ho T-C, Chen M-S, Huang J-S, Yang C-M. Formation of idiopathic macular hole—reappraisal. Graefe’s Archive for Clinical and Experimental Ophthalmology. 2010/06/01 2010;248(6):793–798. doi:10.1007/s00417-009-1297-x

2. Steel DHW, Lotery AJ. Idiopathic vitreomacular traction and macular hole: a comprehensive review of pathophysiology, diagnosis, and treatment. Eye. 2013/10/01 2013;27(1):S1–S21. doi:10.1038/eye.2013.212

3. McCannel CA, Ensminger JL, Diehl NN, Hodge DN. Population-based incidence of macular holes. Ophthalmology. Jul 2009;116(7):1366–9. doi:10.1016/j.ophtha.2009.01.052

4. Lewis ML, Cohen SM, Smiddy WE, Gass JD. Bilaterality of idiopathic macular holes. Graefes Arch Clin Exp Ophthalmol. Apr 1996;234(4):241–5. doi:10.1007/bf00430416

5. Berton M, Robins J, Frigo AC, Wong R. Rate of progression of idiopathic full-thickness macular holes before surgery. Eye (Lond). Aug 2020;34(8):1386–1391. doi:10.1038/s41433-019-0654-1

6. Bringmann A, Unterlauft JD, Barth T, Wiedemann R, Rehak M, Wiedemann P. Different modes of full-thickness macular hole formation. Exp Eye Res. Jan 2021;202:108393. doi:10.1016/j.exer.2020.108393

7. Gass JD. Reappraisal of biomicroscopic classification of stages of development of a macular hole. Am J Ophthalmol. Jun 1995;119(6):752–9. doi:10.1016/s0002-9394(14)72781-3

8. Gass JD. Idiopathic senile macular hole: its early stages and pathogenesis. 1988. Retina. Dec 2003;23(6 Suppl):629–39.

9. Azzolini C, Patelli F, Brancato R. Correlation between optical coherence tomography data and biomicroscopic interpretation of idiopathic macular hole. Am J Ophthalmol. Sep 2001;132(3):348–55. doi:10.1016/s0002-9394(01)01005-4

10. Spaide RF, Wong D, Fisher Y, Goldbaum M. Correlation of vitreous attachment and foveal deformation in early macular hole states. Am J Ophthalmol. Feb 2002;133(2):226–9. doi:10.1016/s0002-9394(01)01377-0

11. Tsai CY, Hsieh YT, Lai TT, Yang CM. Idiopathic macular holes and direction of vitreomacular traction: structural changes and surgical outcomes. Eye (Lond). Dec 2017;31(12):1689–1696. doi:10.1038/eye.2017.141

12. Tsai CY, Hsieh YT, Yang CM. EPIRETINAL MEMBRANE-INDUCED FULL-THICKNESS MACULAR HOLES: The Clinical Features and Surgical Outcomes. Retina. Sep 2016;36(9):1679–87. doi:10.1097/iae.0000000000000999

13. Ang AC, Hsieh YT, Tsui MC, Lai TT, Yang CM. Idiopathic Macular Hole without Vitreomacular Separation. Ophthalmologica. 2022;245(2):187–193. doi:10.1159/000521731

14. Ma IH, Yang CM, Hsieh YT. Wide-based foveal pit: a predisposition to idiopathic epiretinal membrane. Graefes Arch Clin Exp Ophthalmol. Aug 2021;259(8):2095–2102. doi:10.1007/s00417-021-05092-5

15. Ma I-H, Yang C-M, Hsieh Y-T. The Correlations of Macular Structure Characteristics With Idiopathic Epiretinal Membrane and Its Sexual Preference—A Matched Comparison Study. Invest Ophthalmol Vis Sci. 2022;63(2):10–10. doi:10.1167/iovs.63.2.10

16. Bronstein MA, Trempe CL, Freeman HM. Fellow eyes of eyes with macular holes. Am J Ophthalmol. Dec 1981;92(6):757–61. doi:10.1016/s0002-9394(14)75625-9

17. Kumagai K, Ogino N, Hangai M, Larson E. Percentage of Fellow Eyes That Develop Full-Thickness Macular Hole in Patients With Unilateral Macular Hole. Arch Ophthalmol. 2012;130(3):393–394. doi:10.1001/archopthalmol.2011.1427

18. Ezra E, Wells JA, Gray RH, et al. Incidence of idiopathic full-thickness macular holes in fellow eyes. A 5-year prospective natural history study. Ophthalmology. Feb 1998;105(2):353–9. doi:10.1016/s0161-6420(98)93562-x

19. Tick S, Rossant F, Ghorbel I, Gaudric A, Sahel J-A, Chaumet-Riffaud P, Paques M. Foveal Shape and Structure in a Normal Population. Invest Ophthalmol Vis Sci. 2011;52(8):5105–5110. doi:10.1167/iovs.10-7005

20. Bradley A, Applegate RA, Zeffren BS, van Heuven WA. Psychophysical measurement of the size and shape of the human foveal avascular zone. Ophthalmic Physiol Opt. Jan 1992;12(1):18–23.

21. Yoshihara N, Sakamoto T, Yamashita T, Yamashita T, Yamakiri K, Sonoda S, Ishibashi T. Wider retinal artery trajectories in eyes with macular hole than in fellow eyes of patients with unilateral idiopathic macular hole. PLoS One. 2015;10(4):e0122876. doi:10.1371/journal.pone.0122876

22. Nowroozzadeh MH, Moshksar S, Azimi A, Rasti A, Sedaghat A. Comparison of retinal vascular arcade trajectory between eyes with an idiopathic macular hole and the healthy fellow eye. Int Ophthalmol. 2022/01/18 2022;doi:10.1007/s10792-022-02221-9

23. Kumagai K, Hangai M, Larson E, Ogino N. Foveal Thickness in Healthy Fellow Eyes of Patients With Unilateral Macular Holes. Am J Ophthalmol. 2013;156(1):140–148. doi:10.1016/j.ajo.2012.11.029

24. Lindtjørn B, Krohn J, Forsaa VA. Optical coherence tomography features and risk of macular hole formation in the fellow eye. BMC Ophthalmol. 2021;21(1):351–351. doi:10.1186/s12886-021-02111-1

25. Etheridge T, Liu Z, Nalbandyan M, et al. Association of Macular Thickness With Age and Age-Related Macular Degeneration in the Carotenoids in Age-Related Eye Disease Study 2 (CAREDS2), An Ancillary Study of the Women’s Health Initiative. Translational Vision Science & Technology. 2021;10(2):39–39. doi:10.1167/tvst.10.2.39

26. Chung YW, Choi MY, Kim J-S, Kwon J-W. The Association between Macular Thickness and Axial Length in Myopic Eyes. Biomed Res Int. 2019;2019:8913582–8913582. doi:10.1155/2019/8913582

27. Adhi M, Aziz S, Muhammad K, Adhi MI. Macular thickness by age and gender in healthy eyes using spectral domain optical coherence tomography. PLoS One. 2012;7(5):e37638. doi:10.1371/journal.pone.0037638

28. Forsaa VA, Lindtjørn B, Kvaløy JT, Frøystein T, Krohn J. Epidemiology and morphology of full-thickness macular holes. Acta Ophthalmol. Jun 2018;96(4):397–404. doi:10.1111/aos.13618

